# Use of causal claims in observational studies: a research on research study

**DOI:** 10.1101/2020.09.17.20194530

**Authors:** Camila Olarte Parra, Lorenzo Bertizzolo, Sara Schroter, Agnès Dechartres, Els Goetghebeur

## Abstract

**Objective:** To evaluate the consistency of causal statements in the abstracts of observational studies published in The BMJ.

**Design:** Research on research study.

**Data source:** All cohort or longitudinal studies describing an exposure-outcome relationship published in The BMJ during 2018. We also had access to the submitted papers and reviewer reports.

Main outcome measures:

Proportion of published research papers with ‘inconsistent’ use of causal language in the abstract. Papers where language was consistently causal or non-causal were classified as ‘consistently causal’ or ‘consistently not causal’, respectively; those where causality may be inferred were classified as ‘suggests causal’. For the ‘inconsistent’ papers, we then compared the published and submitted version.

**Results:** Of 151 published research papers, 60 described eligible studies. Of these 60, we classified the causal language used as ‘consistently causal’ (13%), ‘suggests causal’ (35%), ‘inconsistent’ (20%) and ‘consistently not causal’(32%). The majority of the ‘Inconsistent’ papers (92%) were already inconsistent on submission. The inconsistencies found in both submitted and published versions was mainly due to mismatches between objectives and conclusions. One section might be carefully phrased in terms of association while the other presented causal language. When identifying only an association, some authors jumped to recommending acting on the findings as if motivated by the evidence presented.

**Conclusion:** Further guidance is necessary for authors on what constitutes a causal statement and how to justify or discuss assumptions involved. Based on screening these abstracts, we provide a list of expressions beyond the obvious ‘cause’ word which may inspire a useful more comprehensive compendium on causal language.

**Strengths and limitations of this study:**

- We present examples of ambiguous causal statements in published abstracts of observational studies in a high impact journal
- We focused on the abstract where clear messages are especially important, as many readers just read the abstract of a study
- The focus on the abstract may miss further discussion on the validity of underlying assumptions justifying causal inference in the setting studied.
- The prevalence and nature of the problems found is a call for better instruction on and consideration of causal language throughout the editorial process in clinical and epidemiological research.
- We provide a list of words and study elements that could point in the direction of causality or otherwise, which may inspire a more comprehensive compendium.

## Introduction

Many researchers remain tempted to draw causal conclusions from observational data despite acknowledging that mere association is not causation because causal inference is the ultimate goal of most clinical and public health research (1, 2). Gold-standard answers are typically sought through randomized controlled trials (RCTs). The unique ability of RCTs to avoid confounding bias (3) has led to demands that empirical research must be drawn from randomized studies to justify causal statements (4-6). RCTs are mainly used to assess the effect of a treatment or intervention but are not easily adapted to evaluate prognostic or risk factors rather than interventions.

There are however good reasons to look beyond RCTs for evidence on treatment effects. In many settings, RCTs are not feasible, ethical or timely and thus observational data are all that is available for some time, as in the recent COVID-19 crisis. Furthermore, observational studies typically involve broader real-world contexts than RCTs, where the costs and risks of experimentation suggest studying high risk patients without major comorbidities (7). This selection challenges generalization to the target population. Trials further suffer from treatment non-compliance which complicates analysis, as treatment-specific populations lose the benefit of randomization. Recent ICH9 guidelines therefore emphasize the importance of causal estimands beyond intention-to-treat, such as per-protocol and as-treated analysis (8, 9).

Deliberately avoiding causal statements on a hoped-for causal answer brings ambiguity and contrived reporting (10, 11). Instead, authors should openly discuss the likely distance in meaning and magnitude between the data based measure they are able to estimate and the desired targeted causal effect. Arguments would consider study design with additional assumptions in context (12). Owing to decades of progress in statistical science (involving potential outcomes, directed acyclic graphs, propensity scores and more) (13), this allows for results, often unreachable by randomized trials, with a justified causal interpretation (14).

In 2010, Cofield et al (5) assessed the use of causal language in observational studies in nutrition but deemed causal language inappropriate for all observational studies. From a different angle, Haber et al (15) examined whether the tone and strength of causal claims made in a given paper matched the language describing the findings in social media. Not surprisingly they found stronger causal statements in the media in half of the cases, emphasizing the importance of clear scientific messages.

To promote this, Lederer et al (16) recently published a guide for authors and editors on how to report causal studies in Respiratory, Sleep and Critical Care Journals. Rather than circumventing the problem by asking to avoid causal language, they provide key elements that ensure valid causal claims (17). Besides briefly explaining causal inference, they provide a definition of a confounder, outline how to identify confounding through so-called directed acyclic graphs and discuss how p-values are often misinterpreted and how their value does not reflect the magnitude, direction or clinical importance of a given association. All these elements empower their target audience to critically assess observational studies.

To find out whether and how statements in study reports present confusing use of causal language (or lack thereof), we examined abstracts of research papers concerned with exposures and outcomes published in The BMJ in 2018. Our focus was on the causal message The BMJ readers receive from the abstracts. We evaluate the consistency of causal statements in the abstracts of observational studies published abstract and if any a priori changes had been made as a result of the peer review process.

## Methods

### Sampling and inclusion criteria

COP identified all original research articles published in The BMJ in 2018 described as either cohort or longitudinal studies in the study design of the abstract. The eligible studies were identified by statements in this section of the abstract such as “cohort”, “longitudinal” or “registry-based”. Those identified as “observational” were included if they suggested a period of follow-up rather than being cross-sectional. Articles described as case-cohorts were excluded as their interpretation and analysis differs from other studies with follow-up assessing the exposure-outcome relationship.

### Assessment of published abstracts

Two reviewers (COP, LB) independently screened all the abstracts of the eligible papers. For the text included under each of the subheadings in the abstract (objective, design, setting, participants, outcome, results, conclusion), the reviewers assessed whether there was an (implicit) causal claim using a yes/no/unclear response. After assessing each separate subheading, each reviewer then gave an overall assessment of the main claims in the paper’s abstract as either ‘consistently causal’, ‘suggests causal’, ‘inconsistent’ or ‘consistently not causal’. After the independent assessments, the overall rating of the abstract was compared between both reviewers; where there was disagreement, a third reviewer (EG) was consulted and a consensus reached.

### Assessment of submitted versions

As the focus of this paper is the avoidance of misleading and ambiguous messages, we further assessed those articles judged as ‘Inconsistent’ to see if there were changes introduced to the manuscript between submission and publication. For this subset, we obtained the submitted version of the manuscripts and the associated peer reviewers’ comments from The BMJ’s manuscript tracking system. We then compared the published version with the first submitted version to identify whether the wording related to causal claims appeared in the submitted version of the abstract and whether changes occurred as a result of comments from peer reviewers and editors.

The same reviewers (COP, LB) independently evaluated the submitted versions of the papers. The reviewers assessed whether the content under each subheading of the submitted abstract differed from the published version. Where there were discrepancies between versions, each reviewer indicated the presence of a causal claim as yes/no/unclear for each abstract subheading (title, objective, design, setting, participants, outcome, results, conclusion) and made an overall assessment of the submitted abstract as either ‘consistently causal’, ‘suggests causal’, ‘inconsistent’ or ‘consistently not causal’. As before, the assessments were compared and, in cases of disagreement, a third reviewer (EG) was consulted and consensus reached.

### Assessment of full text

For the published papers classified as ‘inconsistent’, we further evaluated the full published text to identify the statistical method applied. We look for statements that would support a causal aim, including confounding adjustment, sources of bias and issues of generalizability.

### Ethics and consent

This study used routinely collected data. When authors and reviewers submit manuscripts and reviews to The BMJ, they are notified that their paper or review may be entered into research projects for quality improvement purposes. COP was given access to The BMJ’s data under a confidentiality agreement.

### Patient and public involvement

Patients were not involved in the design, analysis or interpretation of the study. Patients were not participants in this study; it was a methodological study (research on research). Patients’ opinions of causal statements and the use of ambiguous language in research papers is important and further work in this area partnered by patients is important.

## Results

### Assessment of published abstracts

In 2018, 151 research papers were published in The BMJ, of which 60 (40%) were eligible for inclusion in our study (Figure 1). We identified 29 studies (48%) using causal language (‘consistently causal’ and ‘suggests causal’). A further twelve (20%) abstracts were considered inconsistent mainly because the objective stated evaluating an association while the conclusion presented a causal finding (9/12) or the opposite (3/12). Finally, there were abstracts that described studies that aim for prediction or reported associations without (implicitly) suggesting that they had a causal nature that were considered consistently not causal (n=19, 32%).

**Figure 1.**
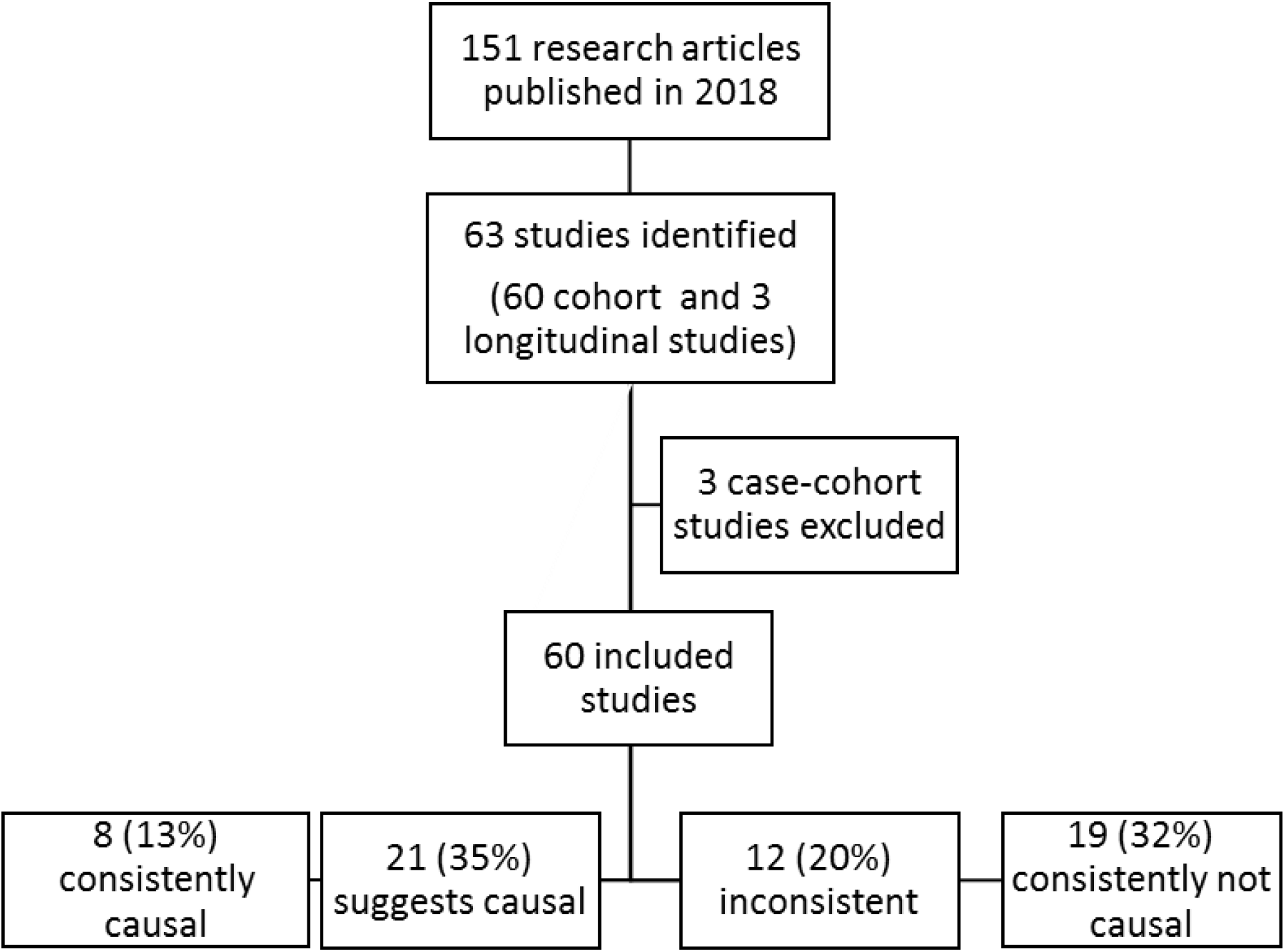
Adapted Prisma flow diagram showing study selection and main study findings.

Table 1 shows excerpts from the abstracts that were evaluated. Each row corresponds to statements from the same study. The first column indicates the assigned category, based on the type of association it describes. The last column explains why a given abstract was considered to belong to the assigned category. As the assessment pertains to causal claims in general, the words referring to the particular topic of the corresponding study were removed from the statements. The examples shown are not an exhaustive list, but were chosen to illustrate the different phrasing of statements belonging to the different categories. It is worth noting that the statements presented correspond to the objective and conclusion subheading of the abstract. When assessing the abstracts, we identified that these were the subheadings under which the information to classify the abstract was mainly found. Other subheadings like Design, Setting and Participants were not as relevant for this purpose.

**Table 1.**
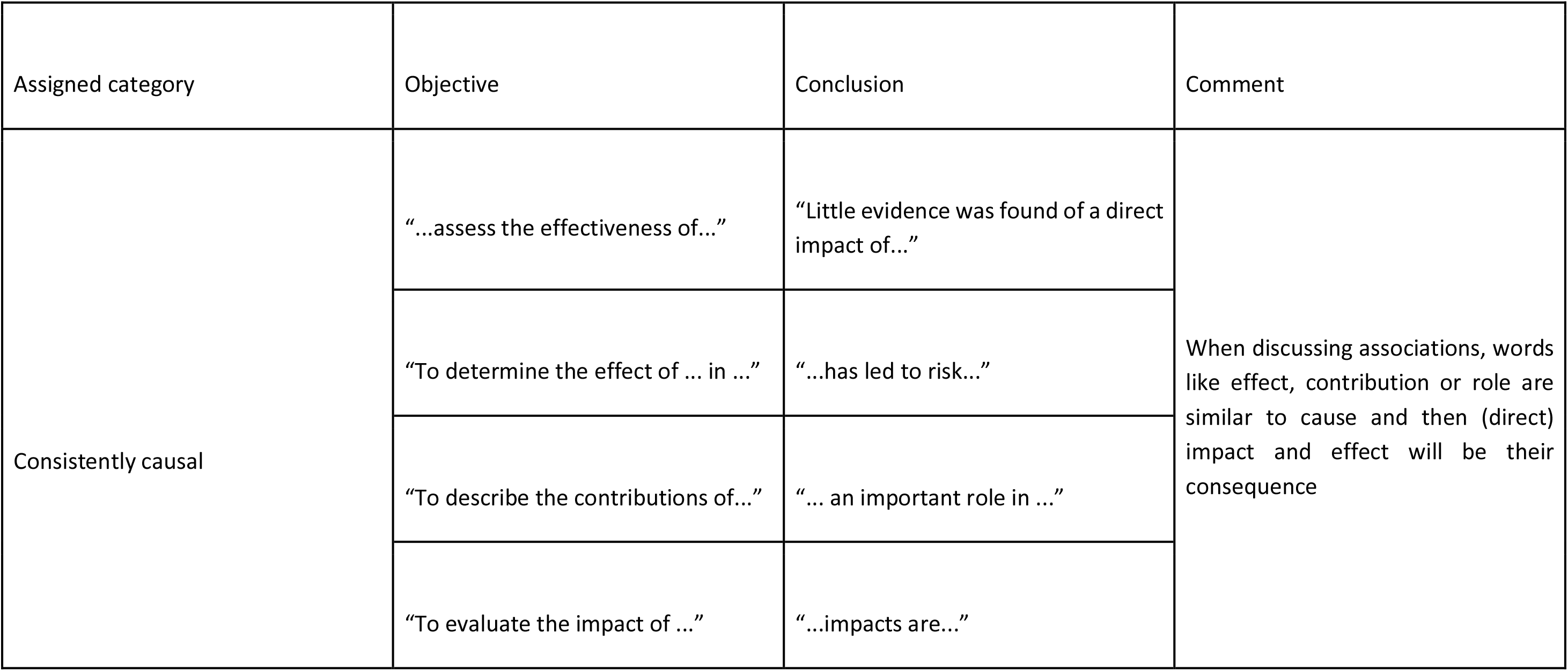
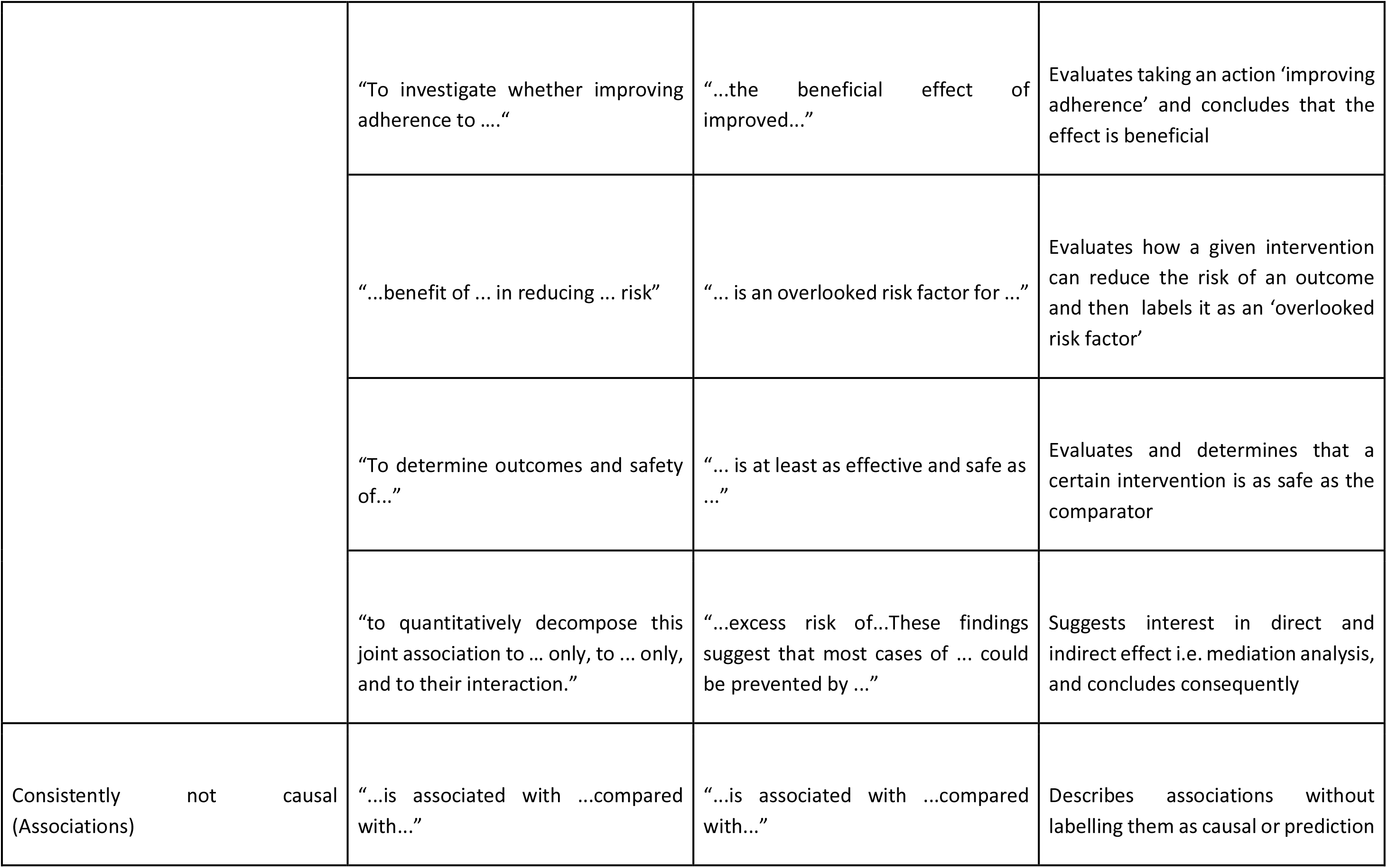
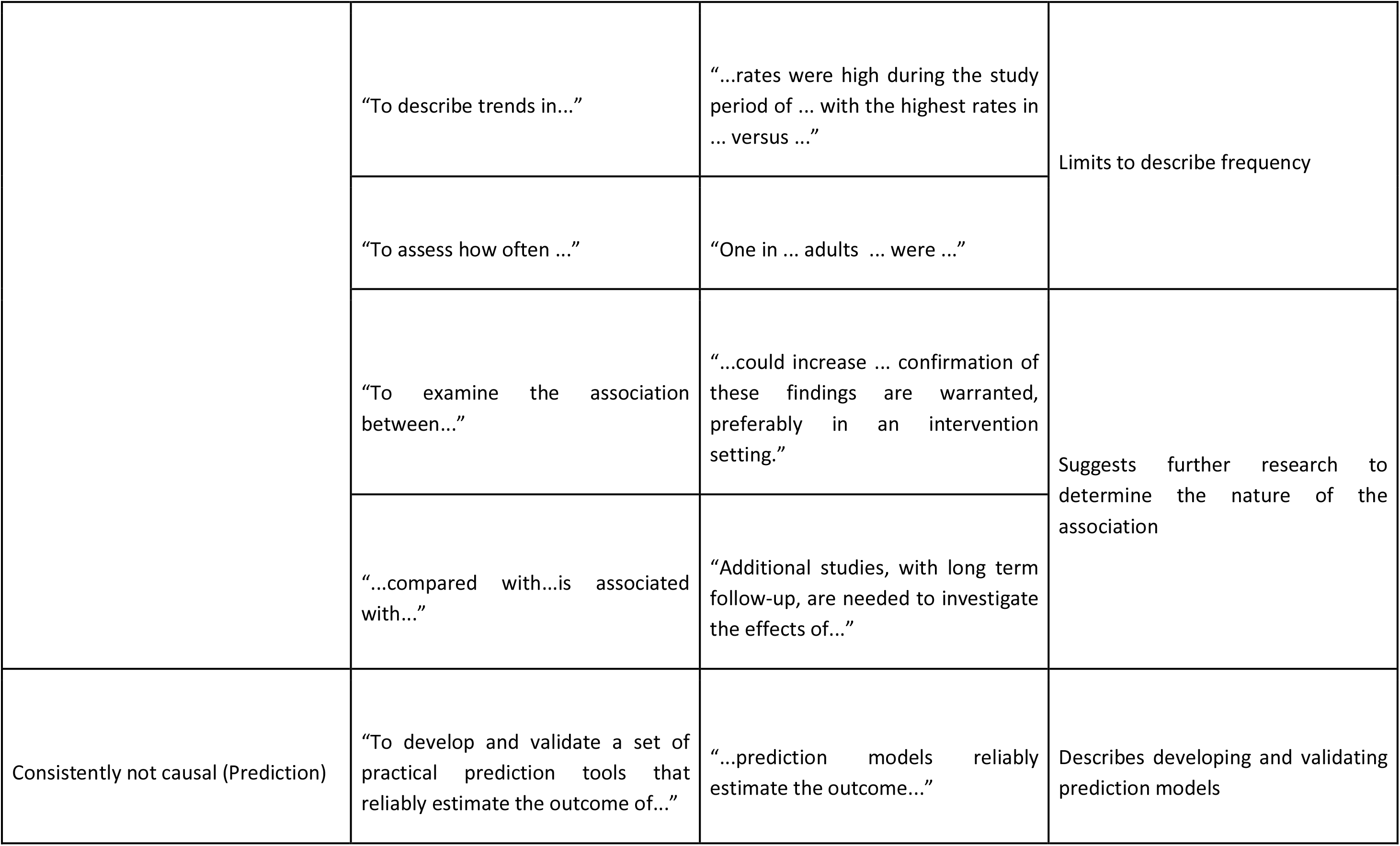
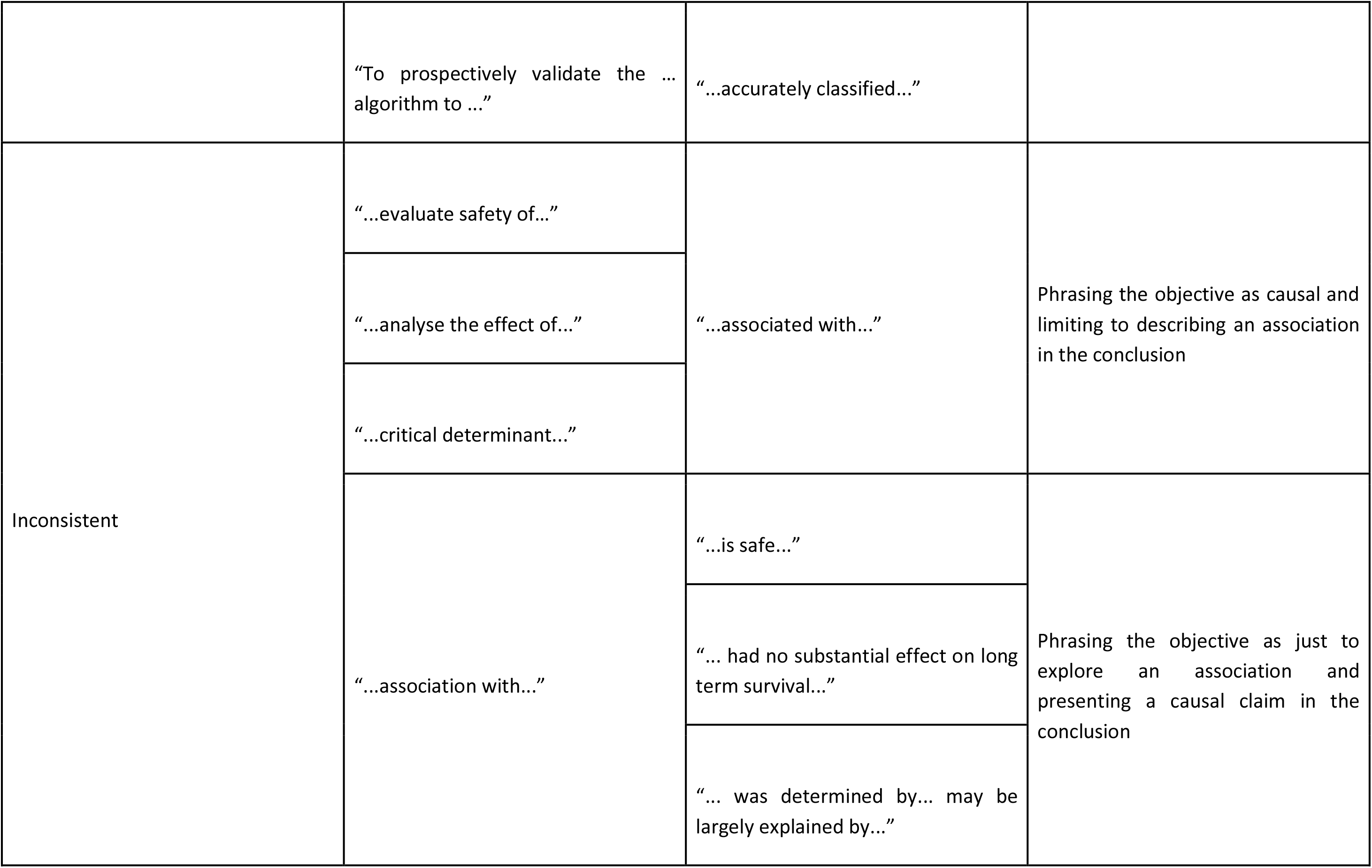
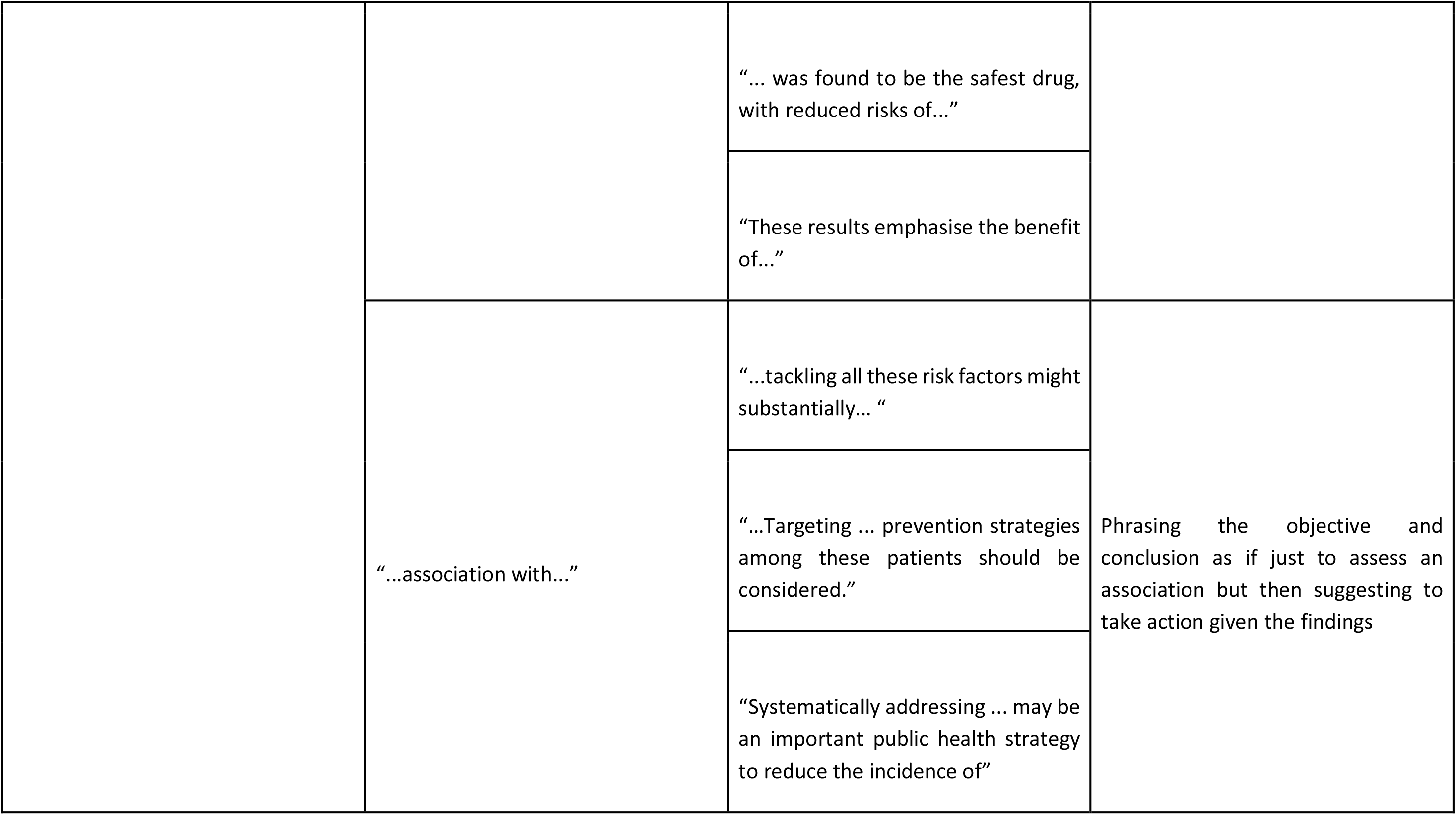
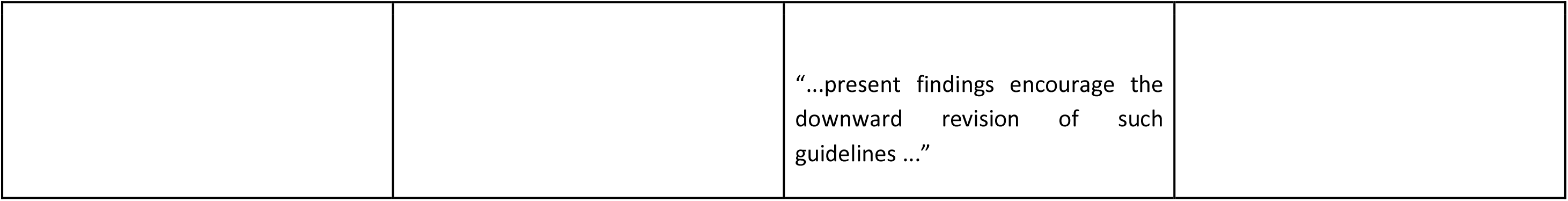
Examples of statements found in the objectives and conclusions of abstracts of observational studies published in The BMJ in 2018 and their corresponding assigned category.

| Assigned category | Objective | Conclusion | Comment |
| --- | --- | --- | --- |
| Consistently causal | "...assess the effectiveness of..." | "Little evidence was found of a direct impact of..." | When discussing associations, words like effect, contribution or role are similar to cause and then (direct) impact and effect will be their consequence |
|  | "To determine the effect of ... in ..." | "...has led to risk..." |  |
|  | "To describe the contributions of..." | "... an important role in ..." |  |
|  | "To evaluate the impact of ..." | "...impacts are..." |  |
|  | “To investigate whether improving adherence to ....” | “...the beneficial effect of improved...” | Evaluates taking an action ‘improving adherence’ and concludes that the effect is beneficial |
|  | “...benefit of ... in reducing ... risk” | “... is an overlooked risk factor for ...” | Evaluates how a given intervention can reduce the risk of an outcome and then labels it as an ‘overlooked risk factor’ |
|  | “To determine outcomes and safety of...” | “... is at least as effective and safe as ...” | Evaluates and determines that a certain intervention is as safe as the comparator |
|  | “to quantitatively decompose this joint association to ... only, to ... only, and to their interaction.” | “...excess risk of...These findings suggest that most cases of ... could be prevented by ...” | Suggests interest in direct and indirect effect i.e. mediation analysis, and concludes consequently |
| Consistently not causal (Associations) | “...is associated with ...compared with...” | “...is associated with ...compared with...” | Describes associations without labelling them as causal or prediction |
|  | "To describe trends in..." | "...rates were high during the study period of ... with the highest rates in ... versus ..." | Limits to describe frequency |
|  | "To assess how often ..." | "One in ... adults ... were ..." |  |
|  | "To examine the association between..." | "...could increase ... confirmation of these findings are warranted, preferably in an intervention setting." | Suggests further research to determine the nature of the association |
|  | "...compared with...is associated with..." | "Additional studies, with long term follow-up, are needed to investigate the effects of..." |  |
| Consistently not causal (Prediction) | "To develop and validate a set of practical prediction tools that reliably estimate the outcome of..." | "...prediction models reliably estimate the outcome..." | Describes developing and validating prediction models |
|  | "To prospectively validate the ... algorithm to ..." | "...accurately classified..." |  |
| Inconsistent | "...evaluate safety of..." | "...associated with..." | Phrasing the objective as causal and limiting to describing an association in the conclusion |
|  | "...analyse the effect of..." |  |  |
|  | "...critical determinant..." |  |  |
|  | "...association with..." | "...is safe..." | Phrasing the objective as just to explore an association and presenting a causal claim in the conclusion |
|  |  | "... had no substantial effect on long term survival..." |  |
|  |  | "... was determined by... may be largely explained by..." |  |
|  |  | “... was found to be the safest drug, with reduced risks of...” |  |
|  |  | “These results emphasise the benefit of...” |  |
|  | “...association with...” | “...tackling all these risk factors might substantially... “ | Phrasing the objective and conclusion as if just to assess an association but then suggesting to take action given the findings |
|  |  | “...Targeting ... prevention strategies among these patients should be considered.” |  |
|  |  | “Systematically addressing ... may be an important public health strategy to reduce the incidence of” |  |
|  |  | “...present findings encourage the downward revision of such guidelines ...” |  |

To further illustrate how statements in these two sections can be misleading, we tabulated a few examples in a 2 by 2 table showing mismatches between what was reported in the objectives and conclusion resulting in the paper being categorised as either ‘consistently (not) causal’ or ‘inconsistent’ (Table 2).

**Table 2.** Examples of (mis)matching causal and non-casual statements found respectively in the objectives and conclusions of abstracts of observational studies published in The BMJ in 2018.

|  |  | Conclusion |  |
| --- | --- | --- | --- |
|  |  | Causal | Not causal |
| Objective | Causal | Consistent<br>“...assess the effectiveness of...” and “Little evidence was found of a direct impact of...”<br>“...benefit of ... in reducing ... risk” and “... is an overlooked risk factor for ...” | Inconsistent<br>“...evaluate safety of...” and “...associated with...”<br>“...analyse the effect of...” and “...associated with...”<br>“...critical determinant...” and “...associated with...” |
|  | Not causal | Inconsistent<br>“...association with...” and “...is safe...”<br>“...association with...” and “... had no substantial effect on long term survival...” | Consistent<br>“To describe trends in...” and “...rates were high during the study period of ... with the highest rates in ... versus ...”<br>“To assess how often ...” and “One in ... adults ... were ...” |
|  |  | “...association with...” and “...tackling all these risk factors might substantially... “<br>“...association with...” and “Systematically addressing ... may be an important public health strategy to reduce the incidence of” | “To develop and validate a set of practical prediction tools that reliably estimate the outcome of...” and “...prediction models reliably estimate the outcome...” |

### Assessment of submitted versions

After evaluating the first submitted version of the 12 abstracts classified as ‘inconsistent’, we classified 11/12 (92%) as also inconsistent on submission. There was only one study where the submitted version described a different type of association. In this case, the conclusion of both the submitted and published versions was rather conservative by stating that the intervention was “independently associated” with the outcome. The submission expressed a causal objective, stating the aim of evaluating the “impact” of a particular intervention with corresponding methods: providing adjusted estimated effects and including sensitivity analysis using propensity score matching. However, in the published version the term “impact” was replaced by “association” making the abstract less clear about a causal aim because both the objectives and the conclusion described an association but the authors still provided adjusted hazard ratios and resorted to propensity score matching.

### Assessment of full text

Looking at the methods used in the papers classified as ‘inconsistent’, we found that 11 of the 12 provided adjusted estimates. Most of the studies (8/12, 67%) used outcome regression models, mainly Cox proportional hazard models, or (propensity score) matching (3/12, 25%). Table 3 presents statements found in both the published abstract and full text of each of these papers regarding the method used and considerations suggesting a causal aim.

**Table 3.** Statements in each of the 12 published observational studies classified as “inconsistent”.

| Abstract |  | Full text |  |  |  |
| --- | --- | --- | --- | --- | --- |
| Objective | Conclusion | Method | Confounder adjustment | Estimates provided | Considerations |
| “To evaluate the relation between [...] and development of [...]” | “ [...] was associated with an increased risk of [...] that was mediated by [...]. Systematically addressing [...] may be an important public health strategy to reduce the incidence of [...] among patients with [...].” | “We calculated the hazard ratios for the relation of [...] to the risk of MRSA using Cox proportional hazard models. [...] We also calculated the absolute risk difference. [...] We performed mediation analyses to examine the extent to which the effect of [...] on the risk of [...] was through [...]. Using marginal structural models we then estimated the natural direct effect [...] and the natural indirect effect [...] while adjusting for the same confounding variables” | “We performed a matched cohort study [...] matched on age (one year either way), sex, and study entry time (within one year either way). Such comparators were chosen to further ensure the comparability [...] In the multivariable Cox model we adjusted for [...].” | “The matched and multivariable adjusted hazard ratios for patients with [...] were 1.69 (1.51 to 1.90) for [...] and 1.26 (1.12 to 1.40) for [...].” | “Our GP practice based dataset could have missed the detection of some inpatient cases of [...]; however, these potential non-differential misclassifications would have biased our results towards the null, rendering our findings conservative” |
| “To quantify the effects of varying [...]” | “[...] is associated with a large increase in [...] among [...] patients. The data from this study suggest that [...] rather than [...] is more strongly associated with [...]” | “For adjusted analysis of time until [...] we used Cox proportional hazards models.” | “Adjusted models included [...]” | “Each additional [...] increased the rate of [...] by 70.7% (95% confidence interval 54.6% to 88.4%) before adjustment and increased the hazard of [...] by 44.0% (40.8% to 47.2%, $P < 0.001$) after adjusting for covariates.” | “To determine the extent to which strong unobserved confounding might explain the observed association, we included this synthetic confounder in a Cox model. [...] As part of a sensitivity analysis, we constructed models that removed potential confounders. [...]” |
| “To evaluate the long term association between [...]” | “Widespread utilisation of [...] may be contributing to long term increased risk of [...]. The potential for [...] should be considered when[...].” | “We conducted Poisson regression analyses using person years as observations.” | “We included several variables as known confounders or effect modifiers in the relation between. [...]The final fully adjusted model adjusted for [...]” | “After adjustment for covariates, the rate ratio was [...] indicating that during the entire period of follow-up the risk of [...] was 21% | “The registered active [...] population is generally representative of the UK population in terms of age, sex, and regional distribution” |
|  |  |  |  | higher during [...] than at other times.” |  |
| “To investigate the association of [...]” | “The shape of the association between [...] and [...] was determined by [...].This finding suggests that the [...] may be largely explained by [...]” | “We used Cox proportional hazards models to estimate hazard ratios and 95% confidence intervals. We stratified the analysis by age in months and calendar year of the questionnaire cycle.” | “For the main analysis, we used [...] measured at baseline to minimize the effect of underlying diseases on mortality [...]. In multivariable models, we adjusted for potential confounders including [...]” | “A multivariable adjusted model showed a positive association between [...] and all cause mortality, whereas [...] showed a U shaped association with all cause mortality. In a mutually adjusted model including both [...] and [...] we consistently observed a strong positive association between [...] and all cause mortality.” | “Our findings remained robust in several sensitivity analyses [...] we cannot entirely rule out the possibility of unmeasured or unknown confounding factors that may account for the associations observed in this study.” |
| “ To examine the associations of [...]” | “This association could be explained by the finding that [...] These results emphasise the importance of revisiting [...] or establishing specific guidelines for management among [...]" | “We performed Cox models with penalised splines” | “In final Cox models with penalised splines, we made adjustments for: [...]” | “[A]fter adjustment for confounding factors, the U shaped association with [...]” |  |
| " To estimate long term survival, health, and educational/social functioning in patients with [...]" | "[...] had no substantial effect on [...]" | "We calculated mortality rate ratios and incidence rate ratios as measures of relative risk." | "For each [...] patient, we used the Danish Civil Registration System and the DNPR to identify all Danish residents with the same sex and date of birth as the patient who had not tested positive [...] and who met the study's inclusion and exclusion criteria [...]. From this population, we extracted 10 people at random for each patient. People in the population comparison cohort were assigned the same date of study inclusion as [...] patients to whom they were matched." | Patients and members of the comparison cohort were well matched with respect to [...] Mortality was not higher among patients in the [...] cohort" |  |
| “To compare the risk of [...]” | “ Although residual confounding cannot be excluded, this finding deserves consideration when [...] is used for [...]” | “We estimated the crude hazard ratio of [...] using Cox proportional hazard regression, and the adjusted hazard ratio was obtained using propensity score matching” | “ We identified potential confounders that were plausibly associated with both [...]based on clinical knowledge [...] In the context of this study, the propensity score is the probability of receiving [...] as opposed to [...], given the baseline characteristics. Patients who received [...] were matched to patients who [...] using a 1:1 nearest neighbor matching algorithm with a caliper of 0.2 of the standard deviation of the propensity score on the logit scale. Covariate balance between the two groups was assessed after matching, and we considered an absolute standardized difference less than 0.1 as evidence of balance” | “The crude hazard ratio of death in the unmatched cohort was 1.51 (95% confidence interval 1.22 to 1.85) and the adjusted hazard ratio in the matched cohort was 1.50 (1.14 to 1.96)” | “Comparison of the baseline characteristics in the unmatched cohort provided little evidence of confounding [...] it is unlikely that a few additional unmeasured variables can explain a 50% increase in the risk independent of all other confounder and proxies of confounders that are adjusted for in our study.” |
| “To determine whether [...] is associated with [...]” | “[...] was independently associated<br>With [...]” | “We fitted both a mixed effect logistic regression model (in which the outcome was defined as dichotomous [...] and the Prentice, Williams, and Peterson (PWP) model” | “We examined the relation between [...] and [...] adjusted for ...” | “The rate of distinct criteria met per year increased by 24% if a patient had been admitted to hospital (hazard ratio 1.24, 95% confidence interval 1.20 to 1.28) when controlled for the other covariates” | “We did a sensitivity analysis using propensity score matching to assess whether the association between [...] and [...] could be due to unmeasured confounders [...] Although we adjusted for a range of characteristics of patients, as with any observational study potential exists for unmeasured confounding, which may partly or fully explain the relation between [...]” |
| “To investigate associations between [...]and to analyse the effect of changes [...]” | “Risks of [...] are inversely associated with [...]” | “We used multivariable Cox regression analysis to compare the rates of [...] and [...].” | “Confounders included in the final models were based on the literature or statistical significance | “Compared with [...], [...] had increased hazard ratios of [...]” | “We believe that our findings are widely applicable and provide justification |
|  |  |  | (P<0.10). The full model included [...]" |  | for [...] and continuing [...]" |
| "To estimate the rates of [...]" | "In cases of [...], approximately [...] will become [...], of which a third will have [...]." | "We present denominators where data for the secondary outcome are missing. We defined the population attributable fraction as $(Re-Run)/Re=(RR-1)/RR$, calculated using Stata. To test the robustness of our findings, we did a sensitivity analysis." | "We compared the demographic and clinical variables of [...]. We used the binomial Wilson score to calculate confidence intervals of single proportions and the Pearson exact method to calculate confidence intervals of risk ratios and medians." | "[...] had a higher risk of [...]The population attributable fraction of [...] was 47% for [...] and 61% for [...]" | "Considering these results when counselling potentially exposed [...] seems reasonable" |
| To perform an expedited assessment of [...] risk associated with exposure to [...]" | "The results do not imply a markedly increased short term overall risk of [...] in [...]." | "We used Cox regression to estimate the hazard ratio with 95% confidence intervals for [...] associated with [...], both for ever use and for the predefined | "Analyses were, however, performed as crude comparisons adjusted only for [...] as well as adjusted for [...] and the potential confounding factors." | "Overall, exposure to [...] showed no association with [...] compared with exposure to [...] (adjusted hazard ratio 1.09, 95% confidence interval 0.85 to 1.41) | "This ensured that the estimates were not affected by immortal time bias [...] As all comparisons were performed within users of [...], the exposure to [...]" |
|  |  | categories of cumulative use” |  | and no evidence of a dose-response relation” | can reasonably be expected to be a random event, and confounding is thus expected to be limited.” |
| “To investigate the risks of [...] in [...]” | “No increased risk of [...] was detected in [...], but increased risks of [...] were found in this study. Our results suggest that [...] risks could be due to [...], rather than [...].” | “To calculate expected [...], we multiplied the person years at risk by corresponding national incidence rates (by 5 year age band and individual calendar year) for the general female population of England and Wales. Standardised incidence ratios were calculated by the comparison of observed values with expected values.” | “We obtained data relating to potential confounding factors such as [...]” | “There was no overall increased risk of [...] (2578 observed v 2641.2 expected [...]; standardised incidence ratio 0.98 (95% confidence interval 0.94 to 1.01); absolute excess risk -2.8 cases per 100 000 person years (95% confidence interval -7.1 to 1.8); table 2)”. | “Given previous inconsistent results, small study size, and lack of information on potential confounders, we undertook a population based linkage study in [...]” |

## Discussion

### Statement of principal findings

We found that the majority of the published research paper abstracts of observational studies had a consistent use of causal language. Still 20% of them contained inconsistent messages on the causal nature of the key “effect”. Inconsistencies showed up in two directions: an intentional quest for causality ending in uncriticized non-causal conclusions or carefully phrased mere associations ending with recommendations to act and intervene based on the exposure outcome association.

Beyond the wording in the abstract, readers can learn much about the sought, after interpretation from described statistical methods, and assumptions made explicit in the paper. On a case by case basis, one could then assess whether additional assumptions, e.g. involving ‘no-unmeasured confounders’, would justify the causal assessment derived from these approaches. Identifying key elements like the ones presented in Table 3 would help to assess if causal inference is possible. If in doubt, a sensitivity analysis may be in order. It seems better to be transparent about the ultimate aim to draw a causal conclusion and to acknowledge to fall short of that, than to generate confusion.

### Comparison with other studies

This is not the first study to evaluate the use of causal language in the medical literature. Cofield et al (5) assessed the use of causal language in observational studies in nutrition. However, they reduced the problem to assessing whether authors included causal language or not, as it was deemed inappropriate due to the observational nature of the study. We have made the case that merely avoiding explicit causal terms is not a real solution. Even without them, a causal conclusion is implicit when the take home message encourages interventions based on the presented findings. Avoiding inconsistency is important but equally one should be able to trust that the use of consistent causal language is not in vain. This requires a more in depth look at methods and assumptions validating the casual claims.

### Strengths and weaknesses of the study

Accurate abstracts are important. In just a few brief paragraphs, the author summarizes key elements of design, methods and results, and comes to a conclusion. Many readers only read the abstract. However, a powerful abstract opens the door to readers and sets the scene for any study. It serves the different roles of informing the audience about its main findings while motivating the reader to further explore the full text, all within the constraints of brevity. This demands from authors special attention to ensure that every word in the abstract is required. All of the above makes the assessment of the abstract relevant but also challenging.

Further research is needed to explore how causal claims presented in the abstract are supported by the full article, which entails assessing the methods used and evaluating whether the underlying assumptions were met (18). The ultimate conclusion should not simply label a study as black or white in causal terms. In the present study we used a convenient limited number of classifications for short statements. In practice a continuous degree of confidence in a potential causal relationship is likely to emerge based on observed association.

We are aware that by limiting our assessment to the abstract, we may have missed the discussion of the extent to which the underlying assumptions that enable causal inference were met. Indeed, when there was a clear causal aim but the authors considered that these assumptions were not fulfilled, they may have decided that a causal claim was inappropriate and phrased their conclusion in terms of association rather than causation. If this is the case, the apparent inconsistency found in the abstract would no longer hold. On the contrary, any undue causal claims can be viewed as a form of spin (19, 20).

### Conclusions and policy implications

As observational data resources abound, methods for causal inference from observational data have surged in tandem with the call for real world evidence. The new opportunities bring new challenges and the responsibility for clear and well supported statements on the evidence. In this spirit and motivated by novel guidelines as proposed by ICH9 and FDA, Miguel Hernan and collaborators have embarked on a project entitled “Developing Guidelines for the Analysis of Randomized Controlled Trials in Real-World Settings”(21). The importance of such initiatives, supports a shift towards being explicit and discussing assumptions underlying causal methods that allow for causal interpretations in context, with or without an RCT (13). In the meantime, uncritical ambiguous phrasing in observational studies remains prevalent (14). Those searching for the best possible evidence supporting future treatment decisions, are best served by transparent reports on observational studies.

Faced with uncertainty when concluding on the nature of the observed exposure outcome relationship, a justifiable balance between the type I and II error rate is a natural guide for action. The cost of errors must be weighed in context, for instance as in clinical trials emphasizing control of the type I error to avoid introducing new unhelpful drugs at a potentially large cost. Alternative weights are typical in screening programs where false positives will be caught in follow-up examinations, but false negatives are lost forever. In a crisis, such as the current COVID-19 pandemic, we must act before long term randomised trials have materialised. It becomes undeniably important to learn as much as we can from observational data, be aware of the types of risk when acting or not, as displayed in the schematic Table 4.

**Table 4.** Impact of the errors of causal effect assignment.

|  |  | True nature of the main exposure effect |  |
| --- | --- | --- | --- |
|  |  | Causal | Not causal |
| Reported nature of the studied exposure effect | Causal | A true causal effect has been discovered. Recommendation to act on this should be considered. Language in the context of a study intended for causal inference. | Type I error: there is no causal effect, but it is claimed.<br>Causal language used or suggestion to take action made when the purpose was to find associations |
|  | Not causal | Type II error: hiding the true causal objective by avoiding use of causal language | No causal language when the objective is prediction or to explore associations |

A prerequisite for good causal language practice includes awareness of which language implies a causal statement and which does not. To support correct phrasing and raise awareness, we have compiled a short list of words and expressions with dedicated (non) causal meaning (Box). The list draws on phrases found in our study and in the references cited, particularly Hernan et al (10) and Thapa et al (6). We consider that a definition of causal language that is generally recognized by the research community is needed (22, 23).

Words like “effect”, “impact”, “determinant of”…, inevitably point in the causal direction and their use should come with the requirement of at least stating and ideally critically evaluating the necessary assumptions (6). Uncertainty on the causal nature of the conclusion should tone down any suggestion for intervening on the studied exposure. Specifying the corresponding level of evidence rather than hiding the ultimate causal aim of a study is what we recommend (19), while acknowledging a margin of error in any empirical study (20).

In summary, we have found that causal messages are embedded in studies otherwise carefully phrased in terms of association. Further guidance for authors appears needed on what constitutes a causal statement, similar to the one published by Lederer et al (13) for Respiratory, Sleep and Critical Care Journals. We look forward to similar guidance for other disease groups. From the screened BMJ abstracts, we provided a list of expressions with clear interpretation which may inspire a useful more comprehensive compendium. We argue that such awareness and special attention amongst authors and reviewers would serve our communication on the best available evidence for conceived interventions.

Box. Examples of words and study elements that could point to causality or otherwise.

|  |  |
| --- | --- |
| Words expressing a causal relationship | • Affect<br>• Attributable<br>• Benefit<br>• Cause<br>• Contribute<br>• Determinant<br>• Efficacy<br>• Impact<br>• Improve<br>• Leads to<br>• Responsible for<br>• Results in<br>• Safety |
| Words that could suggest causality in a given context | • Effect<br>• Induce<br>• Higher (lower) probability<br>• Modify<br>• Risk (factor) |
| Specific expressions avoiding suggestions of casual effects | • Association<br>• Correlation<br>• Less (more) likely link<br>• Predict<br>• Pattern |
| Key aspects suggesting causal aim | • Adjusting for confounders<br>• Discussing 'unmeasured confounders' assumption<br>• Suggesting/recommending intervention<br>• Using instrumental variables<br>• Propensity score adjustment |

## Data Availability

The reviews and published versions of the papers included in the study are publicly available at BMJ.com. No further data will be made available as it is confidential submission data.

## Contributorship statement

- Conceptualization: COP, LB, AD, SS, EG
- Methodology: COP, EG
- Data curation: COP, LB
- Formal analysis: COP, LB, EG
- Writing original draft: COP
- Writing-review and editing: LB, SS, AD, EG
- Approved final version: COP, LB, AD, SS, EG

## Ethics Approval

This study used routinely collected data. When authors and reviewers submit manuscripts and reviews to The BMJ, they are notified that their paper or review may be entered into research projects for quality improvement purposes.

## Transparency declaration

COP confirms that this manuscript is an honest, accurate, and transparent account of the study being reported and that no important aspects of the study have been omitted.

## References

1. Begg MD, March D. Cause and association: Missing the forest for the trees. American Journal of Public Health. 2018;108(5):620-.

2. Hernán MA. A definition of causal effect for epidemiological research. Journal of Epidemiology and Community Health. 2004;58(4):265–71.

3. Kunz R, Oxman aD. The unpredictability paradox: review of empirical comparisons of randomised and non-randomised clinical trials. Bmj. 1998;317(October):1185-90.

4. Ruich P. The use of cause-and-effect language in the JAMA Network journals. 2017.

5. Cofield SS, Corona RV, Allison DB. Use of causal language in observational studies of obesity and nutrition. Obesity Facts. 2010;3(6):353–6.

6. Thapa DK, Visentin DC, Hunt GE, Watson R, Cleary M. Being honest with causal language in writing for publication. Journal of Advanced Nursing. 2020;76(6):1285–8.

7. Pinsky PF, Miller A, Kramer BS, Church T, Reding D, Prorok P, et al. Evidence of a healthy volunteer effect in the prostate, lung, colorectal, and ovarian cancer screening trial. American journal of epidemiology. 2007;165:874-81.

8. Food Drug Administration (FDA), Permutt T, Scott J. E9(R1) Statistical Principles for Clinical Trials: Addendum: Estimands and Sensitivity Analysis in Clinical Trials. 2017;9.

9. European Medicine Agency (EMA). ICH E9 (R1) addendum on estimands and sensitivity analysis in clinical trials to the guideline on statistical principles for clinical trials 2020 17 February 2020

10. Hernan MA. The C-Word: Scientific Euphemisms Do Not Improve Causal Inference From Observational Data. 2018;108(5):616–9.

11. Glymour MM, Hamad R. Causal thinking as a critical tool for eliminating social inequalities in health. American Journal of Public Health. 2018;108(5):623-.

12. Green MJ. Calculating Versus Estimating Causal Effects. American Journal of Public Health. 2018;108(8):e4-e5.

13 Hernán M, Robins J. Causal Inference: What If: Boca Raton: Chapman & Hall/CRC; 2020.

14. Goetghebeur E, Cessie SI, De Stavola B, Moodie E, Waernbaum I. Formulating causal questions and principled statistical answers. 2019.

15. Haber N, Smith ER, Moscoe E, Andrews K, Audy R, Bell W, et al. Causal language and strength of inference in academic and media articles shared in social media (CLAIMS): A systematic review. PLoS ONE. 2018;13(5):1–21.

16. Lederer DJ, Bell SC, Branson RD, Chalmers JD, Marshall R, Maslove DM, et al. Control of Confounding and Reporting of Results in Causal Inference Studies. Guidance for Authors from Editors of Respiratory, Sleep, and Critical Care Journals. Annals of the American Thoracic Society. 2019;16(1):22–8.

17. Harhay MO, Au DH, Dell SD, Gould MK, Redline S, Ryerson CJ, et al. Methodologic Guidance and Expectations for the Development and Reporting of Prediction Models and Causal Inference Studies. Annals of the American Thoracic Society. 2020;17(6):679–82.

18. Hernán MA, Robins JM. Estimating causal effects from epidemiological data. Journal of Epidemiology and Community Health. 2006;60(7):578–86.

19. Boutron I. Reporting and Interpretation of Randomized Controlled Trials With Statistically Nonsignificant Results for Primary Outcomes. JAMA. 2010;303(20):2058-.

20. Ghannad M, Olsen M, Boutron I, Bossuyt PM. A systematic review finds that spin or interpretation bias is abundant in evaluations of ovarian cancer biomarkers. Journal of Clinical Epidemiology. 2019;116:9-17.

21. Developing Guidelines for the Analysis of Randomized Controlled Trials in Real-World Settings [Available from: https://www.pcori.org/research-results/2015/developing-guidelines-analysis-randomized-controlled-trials-real-world.

22. Doan S, Yang EW, Tilak SS, Li PW, Zisook DS, Torii M. Extracting health-related causality from twitter messages using natural language processing. BMC Medical Informatics and Decision Making. 2019;19(Suppl 3).

23. Chiolero A. Causality in public health: One word is not enough. American Journal of Public Health. 2019;109(10):1319–20.

